# Prognostic accuracy of triage tools for adults with suspected COVID-19 in a middle-income setting: an observational cohort study

**DOI:** 10.1101/2022.08.23.22279112

**Authors:** Carl Marincowitz, Laura Sbaffi, Peter Hodkinson, David McAlpine, Gordon Fuller, Steve Goodacre, Peter A. Bath, Madina Hasan, Yasein Omer, Lee Wallis

## Abstract

**Study Objective:** Tools proposed to triage acuity in suspected COVID-19 in the ED have been derived and validated in higher-income settings during early waves of the pandemic. We estimated the accuracy of seven risk-stratification tools recommended to predict severe illness in the Western Cape, South Africa.

**Methods:** An observational cohort study using routinely collected data from EDs across the Western Cape, from the 27^th of^ August 2020 to 11^th^ March 2022 was conducted to assess performance of the PRIEST tool, NEWS2, TEWS, the WHO algorithm, CRB-65, Quick COVID-19 Severity Index and PMEWS in suspected COVID-19. The primary outcome was death or ICU admission.

**Results:** Of 446,084 patients, 15,397 patients (3.45%, 95% CI:34% to 35.1%) experienced the primary outcome. Clinical decision-making for inpatient admission achieved a sensitivity of 0.77 (95% CI 0.76 to 0.78), specificity 0.88 (95% CI 0.87 to 0.88) and the negative predictive value (NPV) 0.99 (95% CI 0.99 to 0.99). NEWS2, PMEWS and PRIEST tool algorithm identified patients at risk of adverse outcomes at recommended cut-offs with moderate sensitivity (>0.8) and specificity ranging from 0.47 (NEWS2) to 0.65 (PRIEST tool). Use of the tools at recommended thresholds would have more than doubled admissions with only a 0.01% reduction in false negative triage.

**Conclusion:** Use of the PRIEST score, NEWS2 and PMEWS at a threshold of a point higher would achieve similar accuracy to current clinical admission decision, with possible gains in transparency and speed of decision-making.

## Background

Mass vaccination and the emergence of the Omicron variant has reduced the severity of illness associated with COVID-19. This has reduced the need for hospitalisation for treatment and the associated risk of emergency health care systems being overwhelmed during periods of increased infection, particularly in high-income settings. However, uneven vaccination in low- and middle-income settings coupled with international relaxation of COVID restrictions and less resilient health care provision mean that emergency health care systems in low- and middle-income countries (LMICs) may still be at risk of being overwhelmed during periods of increased infection.^1, 2^ Although, for now, the acute surge and demands of COVID on health care facilities has relaxed, there is a pressing need for tools to assist clinician decision making, as well as the ability to adapt such tools rapidly to future COVID19 variants or other new epidemics.

In LMICs, disposition decision making is often based on clinician experience and gestalt.^3^ Use of risk-stratification tools to allow rapid triage of need for hospitalisation can help prevent hospitals being overwhelmed and assist less experienced clinicians in ensuring those at risk of deterioration receive inpatient treatment (or at least fair resource allocation). For application in low- and middle-income settings, as rapid COVID tests are not available, triage tools must be applied to patients with suspected COVID and must use easily obtainable clinical factors, as opposed to laboratory and other investigations.^1, 4^ Prognostic research to date has largely been conducted in inpatient and high-income settings on patients with confirmed COVID-19.^4-6^

Acuity scores, including the UK Royal College of Physicians National Early Warning Score, version 2 (NEWS2), COVID specific Pandemic Respiratory Infection Emergency System Triage (PRIEST) score and Quick COVID-19 Severity Index have shown good prediction of adverse outcomes and been suggested as a way to risk-stratify patients with suspected COVID-19 in the Emergency Department.^7-10^ The Triage Early Warning Score (TEWS) is similar to NEWS2 and is used as part of the South African Triage Scale (SATS) in emergency care.^11, 12^ The WHO decision-making algorithm for respiratory infection and CRB-65 are used to risk-stratify patients with bacterial pneumonia and Pandemic Medical Early Warning Score (PMEWS) for use in patients with influenza.^13-15^ However, validation of the accuracy of such risk-stratification tools in low- and middle-income settings in patients with suspected COVID-19 has been limited, as has validation in the Omicron wave.^4^

### Our study aimed to

1. Estimate the accuracy of available risk-stratification tools to predict severe illness in adults with suspected COVID-19 infection in the Western Cape Province of South Africa (a middle-income setting).
2. Assess the accuracy of risk-stratification tools during the Omicron wave.

## Methods

### Study Design

This observational cohort study used routinely collected clinical data from Emergency Departments (EDs) across the Western Cape, from the Hospital Emergency Centre Triage and Information System (HECTIS)^16^ data repository to assess the accuracy in the ED of seven clinical risk-stratification tools (PRIEST tool, Quick Covid-19 Severity Index, TEWS, NEWS2, WHO algorithm, CRB-65 and PMEWS) recommended for use in hospitalised patients with COVID-19 or similar respiratory infections (triage tools shown in Supplementary Material 1).^7, 10, 12, 13, 17-19^ The study was conducted and reported in accordance with Reporting of studies Conducted using Observational Routinely-collected Data (RECORD) guidelines.^20^

### Setting

We obtained data from patients with suspected COVID-19 infection who attended public-sector EDs in the Western Cape Province. This is one of nine provinces in South Africa, and has almost 7 million inhabitants, of whom three quarters use public-sector services.^21^ A convenience sample (based on those hospitals using the recently implemented HECTIS system) was selected of seven hospital EDs, representing predominantly urban, Cape Town metropole district and regional hospitals, as well as a large peri-rural hospital ED. Clinical decision making around COVID patients was largely based on clinician gestalt and experience, contextualised to the local and hospital status, i.e., at times hospitals were overwhelmed with COVID admissions and admission thresholds raised.^22, 23^ Although there were ICU admission tools developed and applied,^24^ there were no specific prognostic or disposition tools applied in the ED beyond routine triage with SATS.

### Data Sources and linkage

Data on ED clinical presentation are routinely collected by the HECTIS system, including presenting complaint, triage variables (using SATS which includes TEWS) and outcome of ED consultation. Through a deterministic matching based on unique patient hospital numbers (performed by the Western Cape Provincial Health Data Centre (PHDC)),^21^ linked data were obtained which included COVID test results from the National Health Laboratory Services (NHLS), comorbidities (based on prior health system encounters, including chronic prescriptions), data around admissions and movements within the health care system during the index COVID encounter, and death (if within, or reported to, the healthcare system). For patients with multiple ED attendances data were extracted from the initial triage data collected for the first ED attendance and outcomes were assessed up to 30 days from this index attendance.

### Inclusion Criteria

Our final cohort consisted of all adults (aged 16 years and over) at time of first (index) ED attendance between 27^th^ August 2020 and 11^th^ March 2022, where a clinical impression of suspected or confirmed COVID-19 infection had been recorded on the HECTIS system. This time period included several waves of COVID19 infection in South Africa, each designated by the responsible variant, comprising the Alpha (March 2020 to September 20220), Beta (November 2020 to February 2021), Delta (May 2021 to October 2021), and Omicron (November 2021 to February 2022).^25^ For those with multiple presentations during the study period, analysis was limited to the index presentation.

### Outcome

The primary composite outcome was intubation or non-invasive ventilation in the ED on index attendance, Intensive Care Unit (ICU) admission or inpatient death up to 30 days from ED index attendance.

The secondary outcomes were inpatient death and ICU admission up to 30 days from index ED attendance.

### Patient Characteristics

Initial physiological parameters and presenting complaints at triage during the patients first (index) presentation to the emergency centre were extracted from the HECTIS database. Where no comorbidities were found, they were assumed not to be present. Implausible physiological variables were set to missing as follows: systolic blood pressure <50 mm HG, temperature >42 or <25 degrees, heart rate < 10/minute, oxygen saturation < 10% and respiratory rate = 0/minute.

### Analysis

We retrospectively applied the 7 triage tools to our cohort to assess their accuracy for the primary and secondary outcomes.^7, 10, 12, 13, 17-19^ Supplementary Material 1 provides details of scoring and handling missing data for the triage tools. For each tool we plotted the receiver operating characteristic (ROC) curve and calculated the area under the ROC curve (c-statistic) for discriminating between patients with and without adverse outcome. We calculated sensitivity, specificity, positive predictive value (PPV) and negative predictive value (NPV) for every threshold of each score for both the primary and secondary outcome. These tools were compared to the sensitivity, specificity, PPV and NPV of decision to admit patients to hospital on index ED attendance. Analysis was conducted for both the whole study population and the subset of patients who presented during the Omicron wave (i.e., patients who presented after the 31^st^ of October 2021).

Calculating performance of each tool at every score allowed different decision-making thresholds to be compared to previously recommended admission thresholds: 0 vs 1+ CRB-65; 0–1 vs 2+ NEWS2; 0–2 vs 3+ PMEWS; 0–4 vs 5+ PRIEST; 0 vs 1 WHO score; TEWS 0-2 vs 2+ Quick COVID-19 Severity Index 0-3 vs 4+. A score of 1 or more for CRB-65 and the WHO score are recommended thresholds for indicating consideration of hospital admission in bacterial pneumonia.^26, 27^ A TEWS score 2+ has previously been proposed as indicating need for inpatient admission.^28^ The NEWS2 and PMEWS thresholds used are lower than previously proposed (0–3 vs 4+ NEWS and 0–3 vs 4+ PMEWS) for triaging patient acuity, and are based on the assessment of their performance in a UK ED population of patients with suspected COVID-19 infection, where higher thresholds gave sub-optimal sensitivity.^8^. In the derivation, the Quick COVID-19 Severity Index a score of 0-3 was recommended as indicating a low risk of progression to respiratory failure.^10^ However, these thresholds were all based on research predominantly conducted in high income settings during early waves of COVID and therefore may not be generalisable to low-middle-income settings or the Omicron variant. All analyses were performed in SPSS version 26.

### Sample Size

The sample size was fixed based on a census sample of patients with suspected COVID in the Western Cape recorded on the HECTIS during the study period.

We a priori assessed the estimated precision of the area under the ROC curve based on a likely 5% event rate for a minimum estimated cohort of 6000 patients. Assuming an AUC of 0.8, based on previous triage tool validation studies,^29^ this sample size would provide an acceptable 95% confidence interval width of 0.06 (95% CI 0.77 to 0.83). In the event the sample size was well in excess of this, and the reported confidence interval width indicates the range in which the population parameter is likely to reside.

### Ethics

The study was approved by the University of Cape Town Human Research Ethics Committee (HREC 594/2021), and the Western Cape Health Research Committee (WC_202111_034). All data were deidentified at source before being provided to the research team.

### Patient and Public Involvement (PPI)

A community advisory board (CAB) was established in advance of the study, comprising eight community members affected by COVID (infected themselves or immediate family infected/ hospitalised). CAB members were recruited by an experienced community liaison officer with links to key community groups. Members were intentionally sought to be representative of the various population groups and demographics of the population. Through several (online) meetings, the CAB were kept abreast of the study, and then given the opportunity to provide input on the outcomes. Participation in the CAB was entirely voluntary with no remuneration beyond expenses incurred.

## Results

### Study population

Figure 1 and Table 1 summarise the study cohort derivation and the characteristics of the 446,084 included patients. In total, 15,397 (3.45%, 95% CI:3.4% to 3.51%) experienced the primary outcome (death, ED intubation/non-invasive ventilation or ICU admission), 11,142 (2.5%, 95% CI: 2.45% to 2.54%) the secondary outcome of death and 2,024 (0.49%, 95% CI: 0.47% to 0.52%) the secondary outcome of ICU admission. The Omicron period included 140,520 patients. Of these, 2,787 (1.98%, 95% CI: 1.91% to 2.06%) experienced the primary outcome, 1,431 (1.02%, 95% CI: 0.97% to 1.07%) the secondary outcome of death and 677 (0.48%, 95% CI: 0.45% to 0.52%) the secondary outcome of ICU admission.

**Table 1.** Patient characteristics by outcome

| Characteristic | Statistic/level | Adverse outcome | No adverse outcome | Total |
| --- | --- | --- | --- | --- |
|  | N | 15,397 (3.45%) | 430,687 (96.55%) | 446,084 |
| Age (years) | Mean (SD) | 55.1 (18) | 43.2 (17.1) | 43.6 (17.2) |
|  | Median (IQR) | 57 (41, 69) | 40 (29, 56) | 41 (29, 57) |
|  | Range | 16 to 105 | 16 to 110 | 16 to 110 |
| Sex | Male | 8,328 (54.1%) | 221,350 (51.4%) | 229,678 (51.5%) |
|  | Female | 7,069 (45.9%) | 209,337 (48.6%) | 216,406 (48.5%) |
| Comorbidities | Asthma/COPD | 2,694 (17.5%) | 63,345 (14.7%) | 66,039 (14.8%) |
|  | Other Chronic respiratory disease | 76 (0.5%) | 946 (0.2%) | 1,022 (0.2%) |
|  | Diabetes | 6,057 (39.3%) | 72,929 (16.9%) | 78,986 (17.7%) |
|  | Hypertension | 6,871 (44.6%) | 116,326 (27%) | 123,197 (27.6%) |
|  | Immunosuppression (HIV) | 2,041 (13.3%) | 75,254 (17.5%) | 77,295 (17.3%) |
|  | Heart Disease | 5,472 (35.5%) | 77,742 (18.1%) | 83,214 (18.7%) |
|  | Pregnant | 105 (0.7%) | 2,642 (0.6%) | 2,747 (0.6%) |
| AVPU | Missing | 652 (4.2%) | 11,826 (2.8%) | 12,478 (2.8%) |
|  | Alert | 10,861 (70.5%) | 389,797 (90.5%) | 400,658 (89.8%) |
|  | Voice | 384 (2.5%) | 5,598 (1.3%) | 5,982 (1.3%) |
|  | Confused | 797 (5.2%) | 16,988 (3.9%) | 17,785 (4%) |
|  | Pain | 794 (5.2%) | 3,177 (0.7%) | 3,971(0.9%) |
|  | Unresponsive | 1,909 (12.4%) | 3,301 (0.8%) | 5,210 (1.2%) |
| Systolic BP (mmHg) | Missing |  |  | 13,784 (3.2%) |
|  | N | 14,489 | 417,499 | 431,988 |
|  | Mean (SD) | 120 (29.5) | 131.2 (25.4) | 131.1 (25.6) |
|  | Median (IQR) | 127 (110,146) | 128 (115,144) | 128 (115,144) |
|  | Range | 50 to 289 | 50 to 300 | 50 to 300 |
| Pulse rate (beats/min) | Missing |  |  | 13,577 (3%) |
|  | N | 14,552 | 417,955 | 432,507 |
|  | Mean (SD) | 99 (23.6) | 93.4 (21) | 93.5 (21.1) |
|  | Median (IQR) | 98 (83,114) | 92 (79,106) | 92 (79,107) |
|  | Range | 11 to 300 | 10 to 300 | 10 to 300 |
| Respiratory rate (breaths/min) | Missing |  |  | 13,540 (3%) |
|  | N | 14,540 | 418,004 | 432,544 |
|  | Mean (SD) | 21.7 (6.5) | 18.5 (4) | 18.6 (4.1) |
|  | Median (IQR) | 20 (18,24) | 18 (16,20) | 18 (16,20) |
|  | Range | 2 to 60 | 1 to 60 | 1 to 60 |
| Oxygen saturation | Missing |  |  | 27,781 (6.2%) |
|  | N | 14,275 | 404,028 | 418,303 |
|  | Mean (SD) | 90.5 (11.5) | 96.3 (5.3) | 96.1 (5.7) |
|  | Median (IQR) | 95 (87,98) | 97 (96,99) | 97 (95,99) |
|  | Range | 10 to 100 | 10 to 100 | 10 to 100 |
| Oxygen administration | Missing |  |  | 26,704 (6%) |
|  | 1 (air) | 7,770 (53.9%) | 377,447 (93.2%) | 385,217 (91.9%) |
|  | 2 (40% O2) | 404 (2.8%) | 7,767 (1.9%) | 8,171 (2%) |
|  | 3 (28% O2) | 10 (0.1%) | 304 (0.08%) | 314 (0.1%) |
|  | 4 (Nasal prongs) | 1,242 (8.6%) | 10,999 (2.7%) | 12,241 (2.9%) |
|  | 5 (FM neb) | 38 (0.3%) | 949 (0.2%) | 987 (0.24%) |
|  | 6 (rebreather mask) | 1,648 (11.4%) | 6,514 (1.6%) | 8,162 (2%) |
|  | 7 (nasal prongs and rebreather mask) | 385 (2.67%) | 996 (0.25%) | 1,381 (0.3%) |
|  | 8 intubated | 2,693 (18.7%) | 0 | 2,693 (0.6%) |
|  | 9 NIV | 218 (1.5%) | 0 | 218 (0.1%) |
| Temperature (°C) | Missing |  |  | 12,510 (2.8%) |
|  | N | 14,743 | 418,831 | 433,574 |
|  | Mean (SD) | 36.4 (1.3) | 36.3 (0.8) | 36.3 (0.8) |
|  | Median (IQR) | 36.4 (35.9,37) | 36.3 (36,36.7) | 36.3 (36,36.7) |
|  | Range | 25 to 41.6 | 25 to 42 | 25 to 42 |
| Cough | Missing |  |  | 135,488 (30.4%) |
|  | Present | 637 (8.2%) | 12,038 (4%) | 12,675 (4.1%) |
| Fever | Missing |  |  | 135,488 (30.4%) |
|  | Present | 203 (2.6%) | 3,998 (1.3%) | 4,201 (1.4%) |
| COVID PCR | Positive | 13,027 (84.6%) | 90,157 (20.9%) | 103,184 (23.1%) |
| Hospital admission | ICU | 2,204 (14.3%) | 0 | 2,204 (0.5%) |
| Death | Within 30 days contact | 11,142 (72.4%) | 0 | 11,142 (2.5%) |

**Figure 1:**
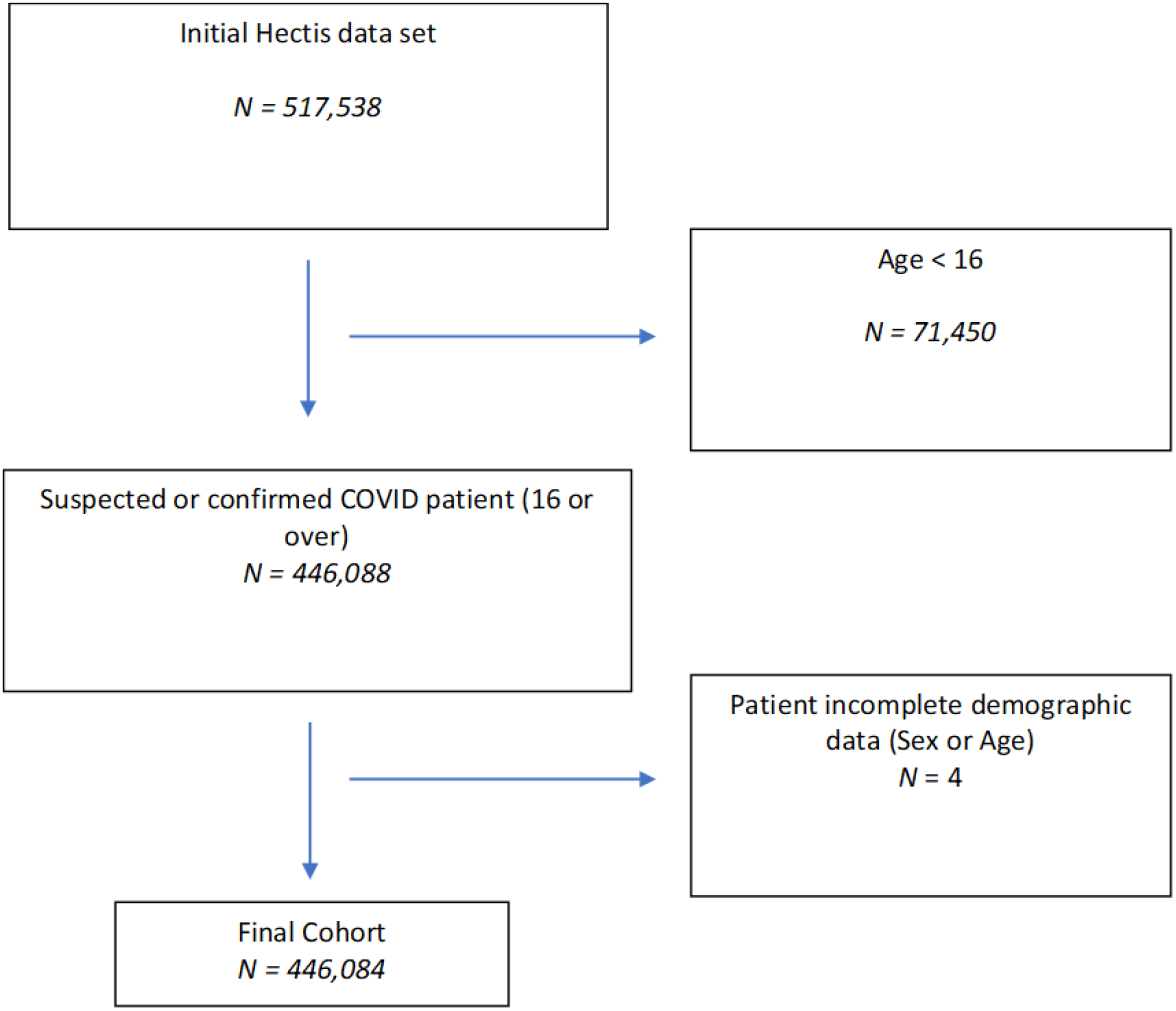
flow diagram of study population selection

In the total cohort of 446,084 patients, 65,657 (14.7%) were admitted as inpatients on index attendance. Of those, 11,862 (18.09%) experienced the primary adverse outcome. Of those not admitted on index attendance, 3,535 (0.9%) experienced the primary outcome. In total, 103,184 patients (23.1%, 95% CI: 23.01 to 23.23%) had a diagnosis of COVID confirmed by PCR testing at a public hospital.

### Triage tool performance

Sensitivity, specificity, positive and negative predictive values for predicting the primary composite outcome using the previously recommended score thresholds are provided in Table 2 and for the Omicron Period in Table 3. Sensitivity and specificity statistics are provided for every score threshold in Supplementary Materials 2 and 3. The ROC curves for these analyses are shown in Figures 2 and 3.

**Table 2.**
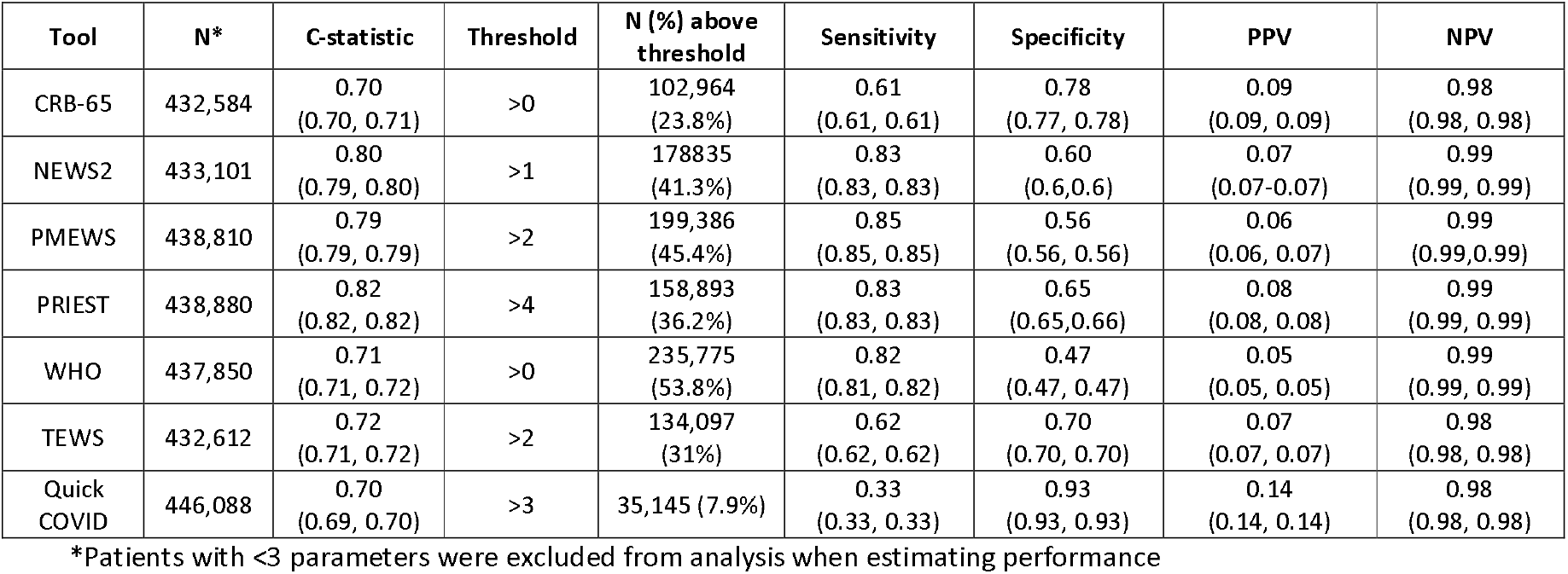
Triage tool diagnostic accuracy statistics (95% CI) for predicting any adverse outcome (entire study period)

| Tool | N* | C-statistic | Threshold | N (%) above threshold | Sensitivity | Specificity | PPV | NPV |
| --- | --- | --- | --- | --- | --- | --- | --- | --- |
| CRB-65 | 432,584 | 0.70<br>(0.70, 0.71) | >0 | 102,964<br>(23.8%) | 0.61<br>(0.61, 0.61) | 0.78<br>(0.77, 0.78) | 0.09<br>(0.09, 0.09) | 0.98<br>(0.98, 0.98) |
| NEWS2 | 433,101 | 0.80<br>(0.79, 0.80) | >1 | 178,835<br>(41.3%) | 0.83<br>(0.83, 0.83) | 0.60<br>(0.6, 0.6) | 0.07<br>(0.07-0.07) | 0.99<br>(0.99, 0.99) |
| PMEWS | 438,810 | 0.79<br>(0.79, 0.79) | >2 | 199,386<br>(45.4%) | 0.85<br>(0.85, 0.85) | 0.56<br>(0.56, 0.56) | 0.06<br>(0.06, 0.07) | 0.99<br>(0.99, 0.99) |
| PRIEST | 438,880 | 0.82<br>(0.82, 0.82) | >4 | 158,893<br>(36.2%) | 0.83<br>(0.83, 0.83) | 0.65<br>(0.65, 0.66) | 0.08<br>(0.08, 0.08) | 0.99<br>(0.99, 0.99) |
| WHO | 437,850 | 0.71<br>(0.71, 0.72) | >0 | 235,775<br>(53.8%) | 0.82<br>(0.81, 0.82) | 0.47<br>(0.47, 0.47) | 0.05<br>(0.05, 0.05) | 0.99<br>(0.99, 0.99) |
| TEWS | 432,612 | 0.72<br>(0.71, 0.72) | >2 | 134,097<br>(31%) | 0.62<br>(0.62, 0.62) | 0.70<br>(0.70, 0.70) | 0.07<br>(0.07, 0.07) | 0.98<br>(0.98, 0.98) |
| Quick COVID | 446,088 | 0.70<br>(0.69, 0.70) | >3 | 35,145 (7.9%) | 0.33<br>(0.33, 0.33) | 0.93<br>(0.93, 0.93) | 0.14<br>(0.14, 0.14) | 0.98<br>(0.98, 0.98) |

\*Patients with <3 parameters were excluded from analysis when estimating performance

**Table 3.** Triage tool diagnostic accuracy statistics (95% CI) for predicting any adverse outcome (Omicron period)

| Tool | N* | C-statistic | Threshold | N (%) above threshold | Sensitivity | Specificity | PPV | NPV |
| --- | --- | --- | --- | --- | --- | --- | --- | --- |
| CRB-65 | 136,961 | 0.69<br>(0.68, 0.70) | >0 | 31,373<br>(22.9%) | 0.59<br>(0.59, 0.59) | 0.78<br>(0.78, 0.78) | 0.05<br>(0.05, 0.05) | 0.99<br>(0.99, 0.99) |
| NEWS2 | 137,125 | 0.77<br>(0.76, 0.78) | >1 | 76,183<br>(55.6%) | 0.87<br>(0.87, 0.87) | 0.45<br>(0.45, 0.45) | 0.03<br>(0.03, 0.03) | 0.99<br>(0.99, 0.99) |
| PMEWS | 138,954 | 0.76<br>(0.75, 0.76) | >2 | 59,876<br>(43.1%) | 0.80<br>(0.80, 0.80) | 0.58<br>(0.58, 0.58) | 0.04<br>(0.04, 0.04) | 0.99<br>(0.99, 0.99) |
| PRIEST | 158,893 | 0.78<br>(0.77, 0.79) | >4 | 46,529<br>(33.5%) | 0.75<br>(0.75, 0.75) | 0.67<br>(0.67, 0.67) | 0.04<br>(0.04, 0.04) | 0.99<br>(0.99, 0.99) |
| WHO | 138,666 | 0.62<br>(0.61, 0.63) | >0 | 72,599<br>(52.4%) | 0.70<br>(0.70, 0.70) | 0.48<br>(0.48, 0.48) | 0.03<br>(0.03, 0.03) | 0.99<br>(0.99, 0.99) |
| TEWS | 136,967 | 0.73<br>(0.72, 0.74) | >2 | 39,509<br>(28.8%) | 0.64<br>(0.64, 0.64) | 0.72<br>(0.72, 0.72) | 0.04<br>(0.04, 0.04) | 0.99<br>(0.99, 0.99) |
| Quick COVID | 140520 | 0.61<br>(0.60, 0.63) | >3 | 8,210<br>(6.4%) | 0.17<br>(0.17, 0.17) | 0.94<br>(0.94, 0.94) | 0.06<br>(0.06, 0.06) | 0.98<br>(0.98, 0.98) |

**Figure 2:**
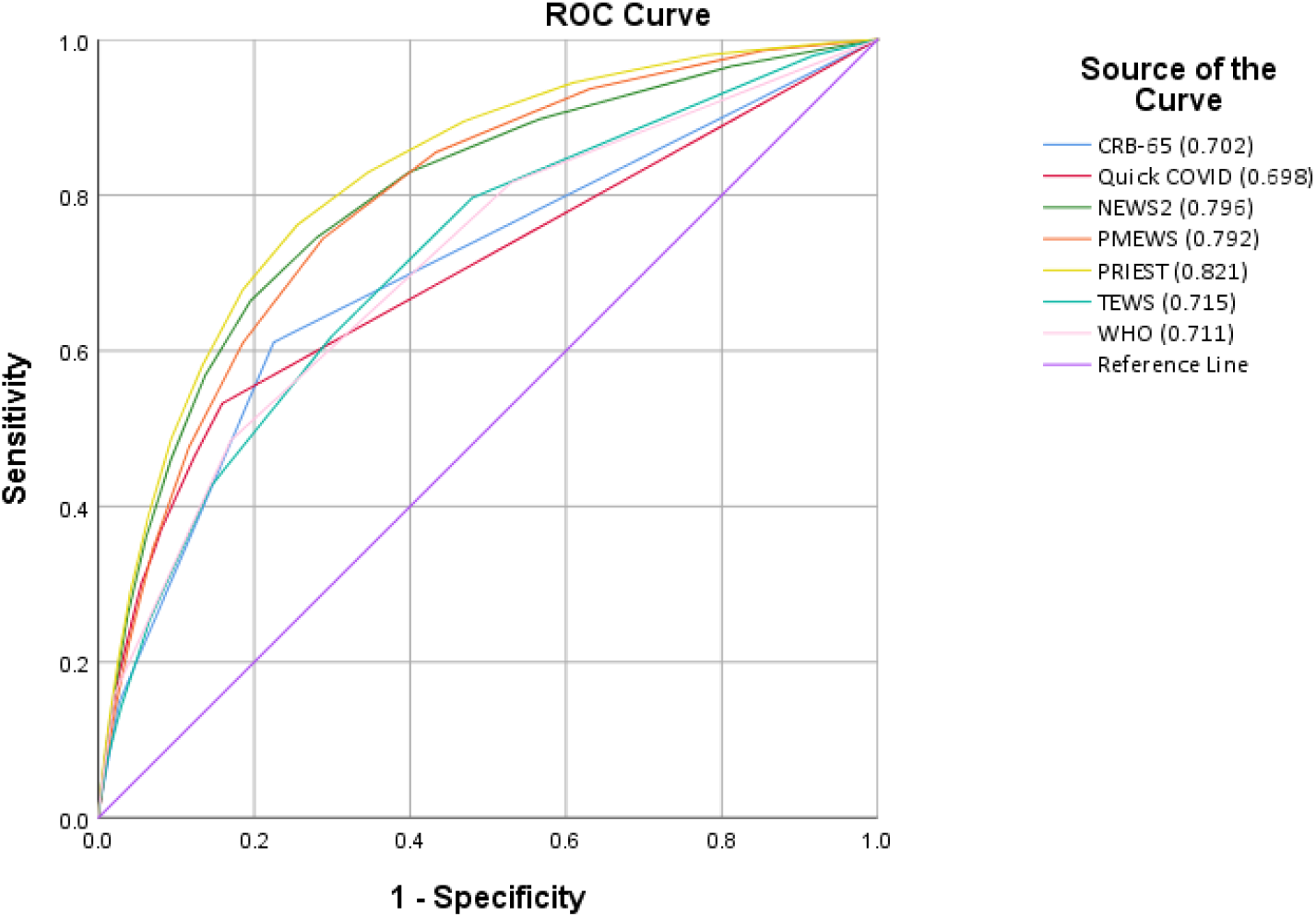
Performance of tools predicting composite primary outcome for total study period

**Figure 3.**
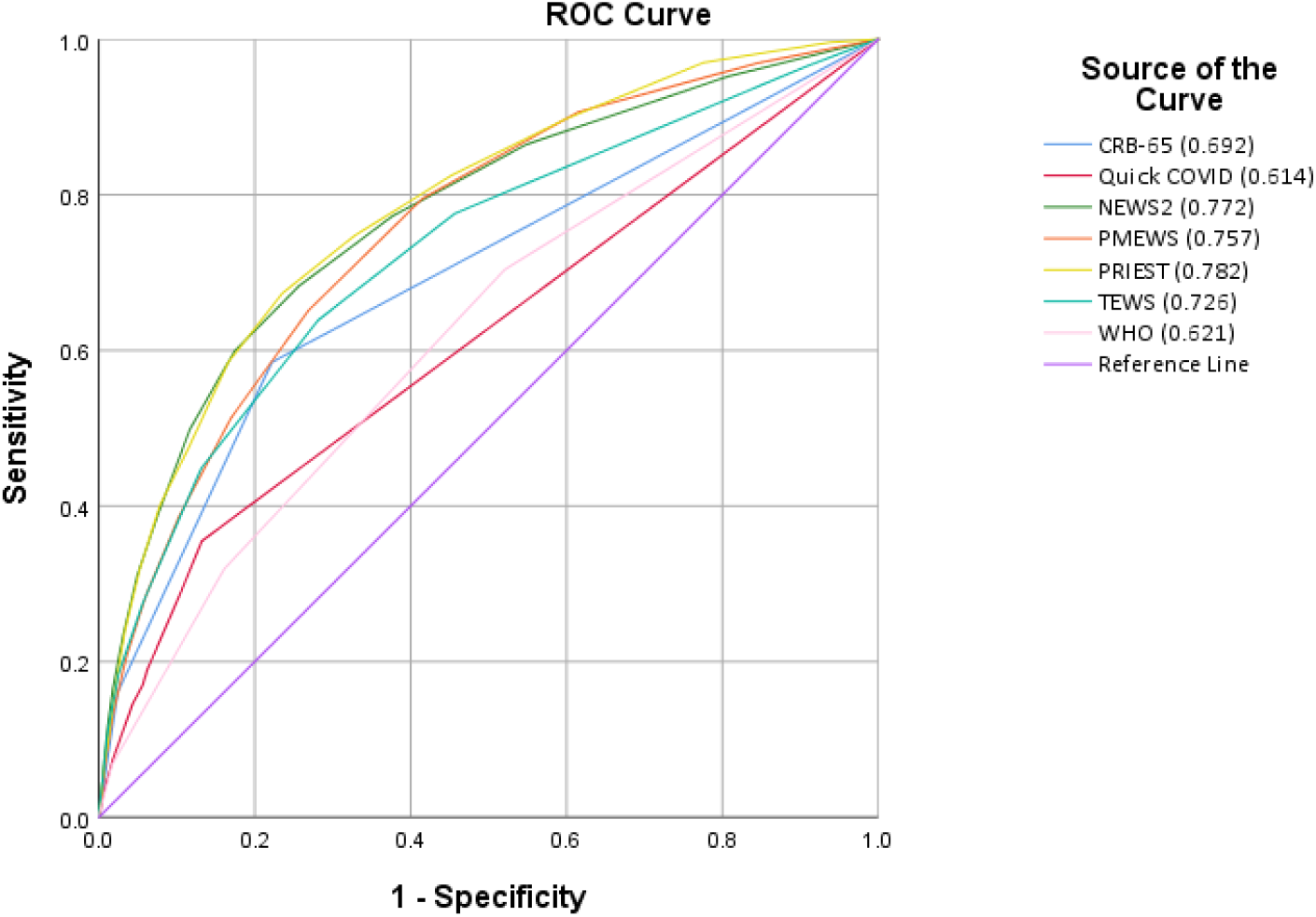
Performance of tools predicting composite primary outcome for the Omicron period

Sensitivity, specificity, positive and negative predictive values for predicting the secondary outcomes (death and ICU admission) using recommended score thresholds are presented in Table 4 and Table 5. The accompanying ROC curves are presented in Supplementary Material 4 and 5.

**Table 4.** Triage tool diagnostic accuracy statistics (95% CI) for predicting death (entire study period)

| Tool | N* | C-statistic | Threshold | N (%) above threshold | Sensitivity | Specificity | PPV | NPV |
| --- | --- | --- | --- | --- | --- | --- | --- | --- |
| CRB-65 | 432,584 | 0.70<br>(0.69, 0.70) | >0 | 102,964<br>(23.8%) | 0.60<br>(0.60, 0.60) | 0.77<br>(0.77, 0.78) | 0.06<br>(0.06, 0.06) | 0.99<br>(0.99, 0.99) |
| NEWS2 | 433,101 | 0.79<br>(0.78, 0.79) | >1 | 178,835<br>(41.3%) | 0.95<br>(0.95, 0.95) | 0.37<br>(0.37, 0.37) | 0.04<br>(0.04, 0.04) | 0.99<br>(0.99, 0.99) |
| PMEWS | 438,810 | 0.81<br>(0.80, 0.81) | >2 | 199,386<br>(45.4%) | 0.88<br>(0.88, 0.88) | 0.56<br>(0.56, 0.56) | 0.05<br>(0.05, 0.05) | 0.99<br>(0.99, 0.99) |
| PRIEST | 438,880 | 0.83<br>(0.83, 0.83) | >4 | 158,893<br>(36.2%) | 0.85<br>(0.85, 0.85) | 0.65<br>(0.65, 0.65) | 0.06<br>(0.06, 0.06) | 0.99<br>(0.99, 0.99) |
| WHO | 437,850 | 0.79<br>(0.79, 0.80) | >0 | 235,775<br>(53.8%) | 0.93<br>(0.93, 0.93) | 0.47<br>(0.47, 0.47) | 0.04<br>(0.04, 0.04) | 0.99<br>(0.99, 0.99) |
| TEWS | 432,612 | 0.70<br>(0.69, 0.70) | >2 | 134,097<br>(31%) | 0.58<br>(0.58, 0.58) | 0.70<br>(0.70, 0.70) | 0.05<br>(0.05, 0.05) | 0.99<br>(0.99, 0.99) |
| Quick COVID | 446,088 | 0.73<br>(0.73, 0.74) | >3 | 35,145 (7.9%) | 0.41<br>(0.41, 0.41) | 0.93<br>(0.93, 0.93) | 0.13<br>(0.13, 0.13) | 0.98<br>(0.98, 0.98) |

\*Patients with <3 parameters were excluded from analysis when estimating performance

**Table 5.** Triage tool diagnostic accuracy statistics (95% CI) for predicting ICU admission (entire study period)

| Tool | N* | C-statistic | Threshold | N (%) above threshold | Sensitivity | Specificity | PPV | NPV |
| --- | --- | --- | --- | --- | --- | --- | --- | --- |
| CRB-65 | 432,584 | 0.57<br>(0.56, 0.59) | >0 | 102,964<br>(23.8%) | 0.38<br>(0.38, 0.38) | 0.76<br>(0.76, 0.76) | 0.008<br>(0.007, 0.009) | 0.99<br>(0.99, 0.99) |
| NEWS2 | 433,101 | 0.72<br>(0.70, 0.73) | >1 | 178,835<br>(41.3%) | 0.83<br>(0.83, 0.83) | 0.43<br>(0.43, 0.43) | 0.007<br>(0.007, 0.007) | 0.99<br>(0.99, 0.99) |
| PMEWS | 438,810 | 0.71<br>(0.70, 0.72) | >2 | 199,386<br>(45.4%) | 0.75<br>(0.75, 0.75) | 0.55<br>(0.55, 0.55) | 0.008<br>(0.008, 0.008) | 0.99<br>(0.99, 0.99) |
| PRIEST | 438,880 | 0.71<br>(0.70, 0.71) | >4 | 158,893<br>(36.2%) | 0.66<br>(0.66, 0.66) | 0.64<br>(0.64, 0.64) | 0.009<br>(0.009, 0.009) | 0.99<br>(0.99, 0.99) |
| WHO | 437,850 | 0.63<br>(0.62, 0.65) | >0 | 235,775<br>(53.8%) | 0.75<br>(0.75, 0.75) | 0.46<br>(0.46, 0.46) | 0.007<br>(0.007, 0.007) | 0.99<br>(0.99, 0.99) |
| TEWS | 432,612 | 0.69<br>(0.67, 0.70) | >2 | 134,097<br>(31%) | 0.59<br>(0.59, 0.59) | 0.69<br>(0.69, 0.69) | 0.009<br>(0.008, 0.009) | 0.99<br>(0.99, 0.99) |
| Quick COVID | 446,088 | 0.65<br>(0.63, 0.66) | >3 | 35,145 (7.9%) | 0.26<br>(0.26, 0.26) | 0.92<br>(0.92, 0.92) | 0.02<br>(0.02, 0.02) | 0.99<br>(0.99, 0.99) |

Clinical decision-making to admit patients to hospital from the ED had a sensitivity of 0.77 (95% CI 0.76 to 0.78) and specificity 0.88 (95% CI 0.87 to 0.88) for the primary outcome. The positive predictive value (PPV) was 0.18 (95% CI 0.18 to 0.18) and the negative predictive value (NPV) 0.99 (95% CI 0.99 to 0.99). Hypothetical use of the PRIEST tool, NEWS2 and PMEWS triage tools would have achieved a higher sensitivity than existing clinical practice to the primary outcome across the study period but this was at a cost of a lower specificity (Table 2). Use of these tools would have more than doubled admissions with only a small reduction in risk of false negative triage. The triage tools generally demonstrated worse discrimination (except TEWS2) during the Omicron period and for ICU admission. The tools all had better discrimination, higher sensitivity but lower specificity when predicting death compared to other outcomes (Table 4).

## Discussion

### Summary

This large retrospective cohort study conducted in a middle-income setting includes data collected from August 2020 to March 2022, encompassing COVID confirmed or clinically suspected COVID patients from the Beta, Delta and Omicron waves in the Western Cape.^25^ The estimated rate of the primary outcome (death, respiratory support or ICU admission) was 3.45% (95% CI:3.4% to 3.51%) across the study period and 1.98% (95% CI: 1.91% to 2.06%) for patients who presented during the Omicron period (31.5% of the cohort).

Existing clinical decision-making only achieved a sensitivity of 0.77 (95% CI 0.76 to 0.78) to the primary outcome, meaning 3, 535 (22.93%) of patients with adverse outcomes were not initially admitted to hospital. However, the low prevalence of the primary outcome in this cohort meant that clinicians could discharge 85.28% of patients on first presentation and discharged patients had less than a 1% chance of experiencing the primary outcome (NPV: 0.99, 95% CI 0.99 to 0.99). Use of the PRIEST, NEWS2 and PMEWS tools at recommended thresholds to inform admission decisions would have improved sensitivity; however, this would have caused a between 21% (PRIEST) and 30% (PMEWS) increase in hospital admissions with only modest associated gains in NPV (Table 2). Potentially using these scores at higher than recommended thresholds could achieve similar admissions with an associated risk of false negative triage similar to current clinical practice (Table 2 and Supplementary Material 6 (risk of adverse outcome each score threshold)).

### Comparison to previous literature

The adverse outcome rate estimated for this study (3.45%) is lower than comparable studies conducted in Europe during the first wave of pandemic.^30, 31^ A UK study reported that 22.1% patients with suspected COVID died or required organ support in an Emergency Department setting.^31^ However, in a mixed cohort of community and hospitalised patients with confirmed COVID from the first wave of the pandemic in China the reported mortality rate was 1.4%.^32^ In European and other high-income settings, the use of telephone triage and other measures may have acted to prevent lower-risk patients with suspected COVID from attending the ED.^33^

The PRIEST tool has been recommended for use by the American College of Emergency Physicians to aid risk-stratification of patients with suspected COVID.^34^ The score has been externally validated in a UK pre-hospital alpha wave cohort and a cohort of 306 patients presenting to EDs in the USA in the winter of 2020-21.^29, 35^ In the development study, the PRIEST score achieved a c-statistic of 0.80 (95%CI 0.79 to 0.81) and, at the recommended threshold of a score above four points, a sensitivity 0.98 (95% 0.97 to 0.98) and specificity 0.34 (95% 0.34 to 0.35) for a composite outcome of death or organ support.^7^ In the American validation study, the score achieved a c-statistic of 0.86 (95% CI: 0.81 to 0.91) and sensitivity of 97.7% (95%CI: 93.2% to 100%) and specificity of 47.2% (95% CI: 41.1% to 53.2%) for a similar outcome^35^. In a UK prehospital study, the PRIEST score achieved a c-statistic of 0.83 (0.82 to 0.84), sensitivity of 0.97 (95%CI: 0.97 to 0.97) and specificity of 0.41 (95%CI: 0.40 to 0.41).^29^

In this study cohort, the PRIEST score (without inclusion of performance status) achieved the best overall discrimination (c-statistic 0.82 (95% CI 0.82 to 0.82)) and sensitivity 0.83 (95% CI 0.83 to 0.83) and specificity of 0.65 (95% CI 0.65 to 0.66) at the recommended threshold to guide hospital admission. Although the discrimination is similar to previous validation studies the differences in accuracy are not explainable solely by the lower prevalence in this study cohort. This may reflect differences in the study population including: younger average age of this study cohort (mean 43.6 years (SD 17.2) versus 62.4 (SD 19.7) in the UK PRIEST cohort) and less impaired physiology (42% of patients NEWS 0 or 1 compared 23% patients in UK PRIEST cohort). As with previous studies, triage tools performed better in this study when predicting death compared to the components of the primary outcome indicating need for organ support (ICU admission in this study).^7, 8, 29^

### Strengths and limitations

This is the first study to use a large cohort of patients identified using routinely collected electronic health care data to validate triage tools in patients with suspected COVID in the Western Cape in the Emergency Department setting. The use of a cohort of patients with suspected infection is important, as this reflects the population which ED staff must clinically triage.^1^ Other prediction models, such as, the International Severe Acute Respiratory Infection Consortium (Coronavirus Clinical Characterisation Consortium) (ISARIC 4C) prediction model requires investigations, including blood tests, and is intended for prediction of inpatient mortality in patients with confirmed COVID, and therefore is not applicable to the rapid triage of need for admission in patients with suspected infection in this setting.^36^ We had low rates of missing data in the variables used in the triage tools assessed (Table 1). Our dataset also comprises multiple COVID waves and allowed the comparison of traige tool performance in the Omicron and earlier waves.^25^

Our evaluation of triage tool accuracy is limited to selected government hospitals in the Western Cape, using the recently implemented HECTIS system and our outcomes are limited to those recorded in hospital. Consequently, deaths are only recorded if the death occurred at a health facility or was specifically notified to a health facility. Deaths at home are not captured, and given the substantial increase in excess deaths attributed to COVID in South Africa (likely at least 68% more for the Western Cape) which occurred undiagnosed at home, it is likely that there were more deaths within 30 days than reflected by the data and the performance of current clinical judgement may be overestimated.^37^ Use of the HECTIS system, electronic records and linking of data from various sources is in its infancy in this context and we are unable to verify the accuracy of individual data, and dependent on a large number of data entry points across facilities and institutions. However, the HECTIS system is used clinically to collect and record the physiological and other variables used to calculate SATS at initial triage in the ED.^11^ Other variables may be recorded less accurately. We assumed that if co-morbidities were not recorded in the routine dataset they were not present. Our cohort is based on the clinical impression of likely COVID infection as determined by the clinical staff performing the initial triage of patients in the ED. This is partly determined by prevalence of COVID-19 infection and clinical guidance which varied during the study period.

### Implications

The PRIEST, NEWS2 and PMEWS triage tools all achieved C-statistics of around 0.8 when estimating death or ICU admission and may show sufficient accuracy to be used clinically in the Western Cape when triaging need for admission in patients with suspected COVID-19. However, in settings with a similarly low prevalence of death or organ support as this study (3.45% compared to 22.1% PRIEST tool development study) use of these tools at previously recommended thresholds would cause large increases in hospital admissions with very small associated gains in reduced risk of false negative triage. The lower risk of death and ICU admission in this study may reflect the role telephone and other pre-hospital triage had in reducing ED attendances of lower-risk patients in the UK and other settings and the lower severity associated with the Omicron wave.^29, 33^ This highlights the need to validate triage tools in different settings and waves to ensure that provide accurate predictions of risk.^3^

Current clinical decision-making to admit patients from the ED in this study cohort achieved a sensitivity of 0.77 (95% CI 0.76 to 0.78) and specificity 0.88 (95% CI 0.87 to 0.88) for the primary outcome. The low prevalence of the primary outcome meant this achieved a negative predictive value (NPV) of 0.99 (95% CI 0.99 to 0.99) and only 14.7% of patients were admitted. Use of the PRIEST score, NEWS2 and PMEWS at a point higher (0–2 vs 3+ NEWS2; 0–3 vs 4+ PMEWS; 0–5 vs 6+ PRIEST) could achieve a similar performance to current clinical practice (Table 2 and Supplementary Material 6). Although, in keeping with previous research,^38^ use of the a triage tool would not outperform clinical decision-making, in LMICs, where disposition decision making is often based on clinician experience and gestalt, use of a triage tool may improve the speed, reproducibility and transparency of decision making, especially for less experienced clinicians.^3^ The PRIEST score, PMEWS and NEWS2 use physiological cut-offs which are not routinely used as part of ED triage using SATS in South Africa and LMICs. The PRIEST score and PMEWS also use predictors, such as performance status, which are not routinely collected in this setting. Development of a triage tool based on existing triage practice and other routinely collected predictive clinical information may improve accuracy and applicability of triage tools in this setting.

The primary outcome used in this study is composite of death and ICU admission (a surrogate for organ support), as this was thought to encompass a definite need for hospital admission.^7^ However, all tools predicted death better than ICU admission (Tables 4 and 5). As has previously been argued, it is important that accuracy of the tools for the primary composite outcome is not used to guide treatment decisions beyond need for admission, such as potential benefit from invasive treatments including ICU admission, as differences in the prediction of death and interventions are likely to mean that the estimation of benefit is inaccurate.^39^

## Conclusion

The NEWS2, PMEWS and PRIEST tools achieved good, estimated discrimination with respect to death or organ support. However, in part due the low prevalence of the primary outcome, use of these tools at previously recommended thresholds would lead to a large increase in hospital admission with a very small associated reduction in false negative triage. Use of the PRIEST score, NEWS2 and PMEWS at a threshold of a point higher would achieve similar accuracy to current clinical admission decision, with possible gains in transparency and speed of decision-making.

## Supporting information

Record Check List

## Data Availability

The data used for this study are subject to data sharing agreements with the Western Cape Department of Health which prohibits further sharing of individual level data. The data sets used are obtainable from this organisation subject to necessary authorisations and approvals

## Authors’ contributions

The idea for the study was conceived by SG, LW, PH, PB, GF and CM. Data processing and linkage was supervised and completed DM. The analyses were completed by LS with additional expert advice from SG and CM. All authors contributed to interpretation of results, read and approved the final manuscript.

## Acknowledgements

This work uses data provided by patients as part of their care and support and the authors wish to recognise Western Cape Department of Health for their contribution of the data that made this research possible, specifically Nesbert Zinyakatira and the team from the Provincial Health Data Centre, Health Impact Assessment Directorate, Western Cape Government Health; the National Health Laboratory Services of South Africa for access to digitized laboratory results; and Dr Moosa Parak and the HECTIS team.

