## Supplementary Material for "Prognostic accuracy of triage tools for adults with suspected COVID-19 in a middle-income setting: an observational cohort study"

**Supplementary Material 1: Triage tool scoring details**

**CRB-65:**

The CRB-65 score is recommended for community settings where access to blood testing is limited. The CRB-65 score uses four parameters, each scoring 1 point when positive and zero if negative, to give a total score between zero and five. It is a 4-point scale that does not include urea with the threshold as <2 for low risk, 2+ for high risk

Four parameters:

1. Confusion: GCS-V is less than 4 or GCS total is less than 15 or AVPU is recorded as V, P or U
2. Respiratory: rate of 30 breaths per minute or more
3. Blood pressure: diastolic BP is 60mmHg or less or systolic BP is 90 mmHg or less
4. Age: 65 years or more

Missing data: If a patient has fewer than three of the five parameters complete, the score was not calculated, otherwise values were assumed to be zero.

**PMEWS:**

PMEWS uses six physiological parameters and patient parameters to calculate a score from zero to 19. The score is calculated by taking the score in the table below dependent on each of the six physiological parameters then adding points for two patient parameters after if they are positive. As living circumstances and performance status were not available in the routine dataset used they were not included in the calculation of the score.

Physiological:

| **Score** | **3** | **2** | **1** | **0** | **1** | **2** | **3** |
| --- | --- | --- | --- | --- | --- | --- | --- |
| **Respiratory Rate** | ≤8 |  |  | 9-18 | 19-25 | 26-29 | ≥30 |
| **SaO₂** | <89 | 90-93 | 94-96 | >96 |  |  |  |
| **Pulse Rate** | ≤40 | 41-50 |  | 51-100 | 101-110 | 111-129 | ≥130 |
| **Systolic BP** | ≤70 | 71-90 | 91-100 | >100 |  |  |  |
| **Temperature** |  | ≤35.0 | 35.1-36.0 | 36.1-37.9 | 38-38.9 | ≥39 |  |
| **Neuro** |  |  |  | Alert | Confused Agitated* | Voice | Pain Uncon |

* confused/agitated will be defined based on GCS-V<4 or GCS total<15

Patient:

1. Add 1 point if age>65
2. Add 1 point if either:
   a. Patient lives alone / no fixed abode or
   b. has a co-morbidity (respiratory, cardiac, renal, immunosuppressed, diabetes)
   c. performance status is more than two suggesting limited activity can self-care, limited activity limited self-care, or bed/chair bound no self-care.

Missing data: If data is missing one or two variables then the normal score (zero) was assumed. If less than three parameters were available the patient was excluded.

**NEWS2:**

The NEWS2 has seven parameters which are scores from zero to three providing an overall score between zero and 20. The scores for each parameter can be found in the table below.

| **Score** | **3** | **2** | **1** | **0** | **1** | **2** | **3** |
| --- | --- | --- | --- | --- | --- | --- | --- |
| **Respiratory Rate** | ≤8 |  | 9-11 | 12-20 |  | 21-24 | ≥25 |
| **SaO₂** | ≤91 | 92-93 | 94-95 | ≥96 |  |  |  |
| **Pulse Rate** | ≤40 |  | 41-50 | 51-90 | 91-110 | 111-130 | ≥131 |
| **Systolic BP** | ≤90 | 91-100 | 101-110 | 111-219 |  |  | ≥220 |
| **Temperature** | ≤35.0 |  | 35.1-36.0 | 36.1-38.0 | 38.1-39.0 | ≥39.1 |  |
| **Neuro** |  |  |  | Alert |  |  | Confusion, Voice, Pain, Unresponsive |
| **Air or Oxygen** |  | Oxygen (based on FiO_2_>21%, or FiO_2_>0 L/min) |  | Air |  |  |  |

Missing data: Any missing data will be imputed with the value zero, therefore classifying missing as normal. The score was not calculated if fewer than three of the parameters were available.

**WHO decision making algorithm for hospitalisation with pneumonia:**

The WHO decision making algorithm for hospitalisation with pneumonia suggests an adult patient is admitted (score 1) if any of the following are present:

- respiratory rate >30/minute,
- oxygen saturation <90%,
- respiratory distress (not included in this evaluation),
- age >60,
- any of the following comorbidities; hypertension, diabetes, cardiovascular disease, chronic respiratory disease, renal impairment or immunosuppression

As a subjective clinical assessment of respiratory distress is not routinely recorded in our data this was not included. Any missing data was assumed as normal. A score was not calculated if fewer than three of the parameters was available.

**The PRIEST clinical severity Score:**

The core PRIEST clinical severity score consists of the seven parameters of NEWS2 and age, sex and performance status. As performances status was not available in the routine data used, this was not included. Scores for each parameter are as follows.

| **Variable** | **Range** | **Score** |
| --- | --- | --- |
| Respiratory rate (per minute) | 12-20 | 0 |
|  | 9-11 | 1 |
|  | 21-24 | 2 |
|  | <9 or >24 | 3 |
| Oxygen saturation (%) | >95 | 0 |
|  | 94-95 | 1 |
|  | 92-93 | 2 |
|  | <92 | 3 |
| Heart rate (per minute) | 51-90 | 0 |
|  | 41-50 or 91-110 | 1 |
|  | 111-130 | 2 |
|  | <41 or >130 | 3 |
| Systolic BP (mmHg) | 111-219 | 0 |
|  | 101-110 | 1 |
|  | 91-100 | 2 |
|  | <91 or >219 | 3 |
| Temperature (C) | 36.1-38.0 | 0 |
|  | 35.1-36.0 or 38.1-39.0 | 1 |
|  | >39.0 | 2 |
|  | <35.1 | 3 |
| Alertness | Alert | 0 |
|  | Confused or not alert | 3 |
| Inspired oxygen | Air | 0 |
|  | Supplemental oxygen | 2 |
| Sex | Female | 0 |
|  | Male | 1 |
| Age (years) | 16-49 | 0 |
|  | 50-65 | 2 |
|  | 66-80 | 3 |
|  | >80 | 4 |
| Performance status | Unrestricted normal activity | 0 |
|  | Limited strenuous activity, can do light activity | 1 |
|  | Limited activity, can self-care | 2 |
|  | Limited self-care | 3 |
|  | Bed/chair bound, no self-care | 4 |

Any missing data was assumed as normal. A score was not calculated if fewer than three of the parameters was available.

**Quick COVID-19 Severity Index (qCSI)**

The qCSI was developed to predict predict respiratory failure within 24 hours of ED attendance as defined by oxygen requirement of greater than 10 L/min by low-flow device, high-flow device, non-invasive or invasive ventilation, or death. The qCSI has three parameters which are scores from zero to five providing an overall score between zero and nine. The scores for each parameter can be found in the table below.

| **Score** | **2** | **1** | **0** | **1** | **2** | **3** | **4** | **5** |
| --- | --- | --- | --- | --- | --- | --- | --- | --- |
| **Respiratory Rate** |  |  | ≥22 | 23-28 | >28 |  |  |  |
| **SaO₂** | ≤88 | 89-92 | >92 |  |  |  |  |  |
| **Oxygen Flow Rate Litres/min** |  |  | ≤2 |  |  |  | 3-4 | >4 |

As it formed part of the original outcome patients on greater than 10L/min oxygen were excluded from analysis in this cohort and patients with missing values were assumed to be in normal range and score zero.

**Triage Early Warning Score (TEWS)**

TEWS is a validated composite triage score which form part of the South African Triage Score, based on judgement of the patient's vital parameters (respiratory rate, heart rate, temperature, systolic blood pressure) and level of consciousness. Possible sum scores range from 0 to 14.

| **Score** | **3** | **2** | **1** | **0** | **1** | **2** | **3** |
| --- | --- | --- | --- | --- | --- | --- | --- |
| **Respiratory Rate** |  | ≤9 |  | 9-14 | 15-20 | 21-29 | ≥29 |
| **Pulse Rate** |  | ≤41 | 41-50 | 51-90 | 91-110 | 111-130 | ≥131 |
| **Systolic BP** | ≤70 | 71-80 | 81-100 | 101-119 |  | ≥199 |  |
| **Temperature** |  | <35 |  | 35-38.4 |  | ≥38.5 |  |
| **Neuro** |  |  |  | Alert | Reacts to Voice | Confusion, reacts to pain | Unresponsive |

Any missing data was assumed as normal. A score was not calculated if fewer than three of the parameters was available

**Supplementary Material 2: Performance of triage tools across the whole range of available scores (overall time period)**

|  |  |  | **Any adverse outcome** | | | |
| --- | --- | --- | --- | --- | --- | --- |
| **Tool** | **Threshold** | **N (%) above threshold** | **Sensitivity (%)** | **Specificity (%)** | **Positive Predictive Value (%)** | **Negative Predictive Value (%)** |
| CRB-65 | >0 | 102964 (23.8) | 61.1 | 77.5 | 8.6 | 98.3 |
|  | >1 | 13135 (3.0) | 14.7 | 97.4 | 16.3 | 97.0 |
|  | >2 | 1035 (0.2) | 1.6 | 99.8 | 22.8 | 96.7 |
|  | >3 | 37 (0.0) | 0.1 | 100.0 | 27.0 | 96.6 |
| NEWS2 | >0 | 355083 (82.0) | 96.7 | 18.5 | 4.0 | 99.4 |
|  | >1 | 249440 (57.6) | 89.8 | 43.5 | 5.3 | 99.2 |
|  | >2 | 178835 (41.3) | 83.0 | 60.2 | 6.8 | 99.0 |
|  | >3 | 128500 (29.7) | 74.7 | 71.9 | 8.5 | 98.8 |
|  | >4 | 91978 (21.2) | 66.7 | 80.4 | 10.6 | 98.6 |
|  | >5 | 65823 (15.2) | 57.0 | 86.3 | 12.7 | 98.3 |
|  | >6 | 45545 (10.5) | 45.9 | 90.7 | 14.8 | 98.0 |
|  | >7 | 30757 (7.1) | 35.9 | 93.9 | 17.1 | 97.7 |
|  | >8 | 19903 (4.6) | 26.2 | 96.2 | 19.3 | 97.4 |
|  | >9 | 12072 (2.8) | 17.2 | 97.7 | 20.9 | 97.1 |
|  | >10 | 7272 (1.7) | 11.1 | 98.6 | 22.3 | 96.9 |
|  | >11 | 4234 (1.0) | 6.9 | 99.2 | 23.9 | 96.8 |
|  | >12 | 2509 (0.6) | 4.5 | 99.6 | 26.2 | 96.7 |
|  | >13 | 1349 (0.3) | 2.5 | 99.8 | 27.7 | 96.7 |
|  | >14 | 722 (0.2) | 1.5 | 99.9 | 30.9 | 96.7 |
|  | >15 | 370 (0.1) | 0.9 | 99.9 | 36.2 | 96.6 |
|  | >16 | 165 (0.0) | 0.4 | 100.0 | 32.1 | 96.6 |
|  | >17 | 58 (0.0) | 0.1 | 100.0 | 32.8 | 96.6 |
|  | >18 | 20 (0.0) | 0.0 | 100.0 | 20.0 | 96.6 |
|  | >19 | 4 (0.0) | 0.0 | 100.0 | 0.0 | 96.6 |
| PMEWS | >0 | 375059 (85.5) | 98.4 | 15.0 | 3.9 | 99.6 |
|  | >1 | 280430 (63.9) | 93.4 | 37.1 | 5.0 | 99.4 |
|  | >2 | 199386 (45.4) | 85.0 | 56.0 | 6.4 | 99.1 |
|  | >3 | 138152 (31.5) | 74.0 | 70.0 | 8.0 | 98.7 |
|  | >4 | 92477 (21.1) | 60.6 | 80.3 | 9.8 | 98.3 |
|  | >5 | 59680 (13.6) | 47.5 | 87.6 | 11.9 | 97.9 |
|  | >6 | 36777 (8.4) | 34.4 | 92.5 | 14.0 | 97.6 |
|  | >7 | 21636 (4.9) | 22.9 | 95.7 | 15.8 | 97.2 |
|  | >8 | 12083 (2.8) | 14.2 | 97.7 | 17.6 | 97.0 |
|  | >9 | 6292 (1.4) | 8.1 | 98.8 | 19.3 | 96.8 |
|  | >10 | 3119 (0.7) | 4.2 | 99.4 | 20.3 | 96.7 |
|  | >11 | 1483 (0.3) | 2.3 | 99.7 | 22.8 | 96.7 |
|  | >12 | 682 (0.2) | 1.1 | 99.9 | 24.3 | 96.6 |
|  | >13 | 309 (0.1) | 0.5 | 99.9 | 25.6 | 96.6 |
|  | >14 | 119 (0.0) | 0.2 | 100.0 | 24.4 | 96.6 |
|  | >15 | 27 (0.0) | 0.0 | 100.0 | 22.2 | 96.6 |
|  | >16 | 4 (0.0) | 0.0 | 100.0 | 50.0 | 96.6 |
| PRIEST | >0 | 413109 (94.1) | 99.5 | 6.1 | 3.6 | 99.7 |
|  | >1 | 343770 (78.3) | 97.9 | 22.4 | 4.3 | 99.7 |
|  | >2 | 271120 (61.8) | 94.4 | 39.4 | 5.2 | 99.5 |
|  | >3 | 210878 (48.0) | 89.3 | 53.4 | 6.3 | 99.3 |
|  | >4 | 158893 (36.2) | 82.8 | 65.4 | 7.8 | 99.1 |
|  | >5 | 119112 (21.1) | 75.8 | 74.6 | 9.5 | 98.9 |
|  | >6 | 88405 (20.1) | 67.4 | 81.5 | 11.4 | 98.6 |
|  | >7 | 64649 (14.7) | 57.5 | 86.8 | 13.3 | 98.3 |
|  | >8 | 46543 (10.6) | 48.2 | 90.7 | 15.5 | 98.0 |
|  | >9 | 32504 (7.4) | 38.1 | 93.7 | 17.5 | 97.7 |
|  | >10 | 21466 (4.9) | 28.4 | 95.9 | 19.7 | 97.4 |
|  | >11 | 13648 (3.1) | 19.8 | 97.5 | 21.7 | 97.2 |
|  | >12 | 8192 (1.9) | 12.8 | 98.5 | 23.4 | 97.0 |
|  | >13 | 5044 (1.1) | 8.7 | 99.1 | 25.7 | 96.9 |
|  | >14 | 3038 (0.7) | 5.4 | 99.5 | 26.8 | 96.8 |
|  | >15 | 1805 (0.4) | 3.4 | 99.7 | 27.8 | 96.7 |
|  | >16 | 1028 (0.2) | 1.9 | 99.8 | 27.6 | 96.7 |
|  | >17 | 534 (0.1) | 1.1 | 99.9 | 29.4 | 96.6 |
|  | >18 | 271 (0.1) | 0.5 | 100.0 | 29.5 | 96.6 |
|  | >19 | 99 (0.0) | 0.2 | 100.0 | 28.3 | 96.6 |
|  | >20 | 43 (0.0) | 0.1 | 100.0 | 27.9 | 96.6 |
|  | >21 | 12 (0.0) | 0.0 | 100.0 | 16.7 | 96.6 |
|  | >22 | 6 (0.0) | 0.0 | 100.0 | 0.0 | 96.6 |
|  | >23 | 2 (0.0) | 0.0 | 100.0 | 0.0 | 96.6 |
|  | >24 | 2 (0.0) | 0.0 | 100.0 | 0.0 | 96.6 |
| WHO | >0 | 235775 (53.8) | 81.5 | 47.1 | 5.1 | 98.6 |
| TEWS | >0 | 397455 (91.9) | 97.9 | 8.3 | 3.6 | 99.1 |
|  | >1 | 212145 (49.0) | 79.7 | 52.0 | 5.5 | 98.7 |
|  | >2 | 134097 (31.0) | 61.9 | 70.1 | 6.7 | 98.1 |
|  | >3 | 66751 (15.4) | 42.7 | 85.5 | 9.3 | 97.7 |
|  | >4 | 31047 (7.2) | 25.3 | 93.5 | 11.9 | 97.3 |
|  | >5 | 14168 (3.3) | 14.1 | 97.1 | 14.5 | 97.0 |
|  | >6 | 6175 (1.4) | 7.7 | 98.8 | 18.2 | 96.8 |
|  | >7 | 2574 (0.6) | 3.6 | 99.5 | 20.3 | 96.7 |
|  | >8 | 948 (0.2) | 1.6 | 99.8 | 25.0 | 96.7 |
|  | >9 | 316 (0.1) | 0.6 | 99.9 | 27.8 | 96.7 |
|  | >10 | 98 (0.0) | 0.2 | 100.0 | 32.7 | 96.6 |
|  | >11 | 32 (0.0) | 0.1 | 100.0 | 37.5 | 96.6 |
|  | >12 | 8 (0.0) | 0.0 | 100.0 | 50.0 | 96.6 |
| Quick COVID | >0 | 74887 (16.8) | 51.0 | 84.4 | 10.5 | 98.0 |
|  | >1 | 58897 (13.2) | 44.6 | 87.9 | 11.6 | 97.8 |
|  | >2 | 39161 (8.8) | 35.3 | 92.2 | 13.9 | 97.6 |
|  | >3 | 35145 (7.9) | 32.8 | 93.0 | 14.3 | 97.5 |
|  | >4 | 27816 (6.2) | 28.9 | 94.6 | 16.0 | 97.4 |
|  | >5 | 13898 (3.1) | 17.5 | 97.4 | 19.4 | 97.1 |
|  | >6 | 5700 (1.3) | 8.6 | 99.0 | 23.2 | 96.8 |
|  | >7 | 1840 (0.4) | 2.3 | 99.7 | 19.5 | 96.6 |
|  | >8 | 1675 (0.4) | 2.2 | 99.7 | 20.7 | 96.6 |
|  | >9 | 721 (0.2) | 1.2 | 99.9 | 25.2 | 96.6 |
|  | >10 | 237 (0.1) | 0.5 | 100.0 | 30.4 | 96.6 |

**Supplementary Material 3: Performance of triage tools across the whole range of available scores (Omicron period)**

|  |  |  | **Any adverse outcome** | | | |
| --- | --- | --- | --- | --- | --- | --- |
| **Tool** | **Threshold** | **N (%) above threshold** | **Sensitivity (%)** | **Specificity (%)** | **Positive Predictive Value (%)** | **Negative Predictive Value (%)** |
| CRB-65 | >0 | 31373 (22.9) | 58.5 | 77.8 | 5.0 | 98.9 |
|  | >1 | 3699 (2.7) | 16.1 | 97.6 | 11.7 | 98.1 |
|  | >2 | 299 (0.2) | 1.3 | 99.8 | 12.0 | 98.1 |
|  | >3 | 12 (0.0) | 0.1 | 100.0 | 33.3 | 98.0 |
| NEWS2 | >0 | 111111 (81.0) | 95.3 | 19.3 | 2.3 | 99.5 |
|  | >1 | 76183 (55.6) | 86.6 | 45.1 | 3.1 | 99.4 |
|  | >2 | 52801 (38.5) | 77.5 | 62.3 | 4.0 | 99.3 |
|  | >3 | 36533 (26.6) | 68.6 | 74.2 | 5.1 | 99.2 |
|  | >4 | 25109 (18.3) | 60.2 | 82.5 | 6.5 | 99.0 |
|  | >5 | 17177 (12.5) | 50.3 | 88.2 | 8.0 | 98.9 |
|  | >6 | 11487 (8.4) | 39.5 | 92.3 | 9.3 | 98.7 |
|  | >7 | 7466 (5.4) | 31.2 | 95.1 | 11.4 | 98.6 |
|  | >8 | 4828 (3.5) | 23.4 | 96.9 | 13.2 | 98.4 |
|  | >9 | 2909 (2.1) | 16.6 | 98.2 | 15.5 | 98.3 |
|  | >10 | 1835 (1.3) | 11.6 | 98.9 | 17.2 | 98.2 |
|  | >11 | 1081 (0.8) | 7.0 | 99.3 | 17.7 | 98.1 |
|  | >12 | 667 (0.5) | 4.2 | 99.8 | 17.1 | 98.1 |
|  | >13 | 346 (0.3) | 2.2 | 99.8 | 17.6 | 98.1 |
|  | >14 | 183 (0.1) | 1.2 | 99.9 | 18.0 | 98.0 |
|  | >15 | 78 (0.1) | 0.8 | 100.0 | 26.9 | 98.0 |
|  | >16 | 44 (0.0) | 0.6 | 100.0 | 34.1 | 98.0 |
|  | >17 | 26 (0.0) | 0.3 | 100.0 | 34.6 | 98.0 |
|  | >18 | 6 (0.0) | 0.1 | 100.0 | 33.3 | 98.0 |
| PMEWS | >0 | 117215 (84.4) | 96.6 | 15.9 | 2.3 | 99.6 |
|  | >1 | 86096 (62.0) | 90.6 | 38.6 | 2.9 | 99.5 |
|  | >2 | 59876 (43.1) | 79.5 | 57.7 | 3.7 | 99.3 |
|  | >3 | 39870 (28.7) | 64.2 | 72.0 | 4.5 | 99.0 |
|  | >4 | 25880 (18.6) | 50.8 | 82.0 | 5.4 | 98.8 |
|  | >5 | 16089 (11.6) | 39.2 | 89.0 | 6.7 | 98.6 |
|  | >6 | 9740 (7.0) | 28.8 | 93.4 | 8.2 | 98.5 |
|  | >7 | 5628 (4.1) | 20.8 | 96.3 | 10.3 | 98.4 |
|  | >8 | 3013 (2.2) | 12.9 | 98.0 | 11.8 | 98.2 |
|  | >9 | 1552 (1.1) | 7.4 | 99.0 | 13.1 | 98.1 |
|  | >10 | 813 (0.6) | 3.9 | 99.5 | 13.3 | 98.1 |
|  | >11 | 377 (0.3) | 1.9 | 99.8 | 14.1 | 98.0 |
|  | >12 | 198 (0.1) | 1.3 | 99.9 | 17.7 | 98.0 |
|  | >13 | 100 (0.1) | 0.6 | 99.9 | 17.0 | 98.0 |
|  | >14 | 36 (0.0) | 0.1 | 100.0 | 11.1 | 98.0 |
|  | >15 | 12 (0.0) | 0.1 | 100.0 | 16.7 | 98.0 |
|  | >16 | 4 (0.0) | 0.1 | 100.0 | 50.0 | 98.0 |
| PRIEST | >0 | 130304 (93.8) | 99.4 | 6.4 | 2.1 | 99.8 |
|  | >1 | 107749 (77.5) | 96.9 | 22.9 | 2.5 | 99.7 |
|  | >2 | 83661 (60.2) | 89.8 | 40.4 | 3.0 | 99.5 |
|  | >3 | 63585 (45.8) | 82.4 | 55.0 | 3.6 | 99.4 |
|  | >4 | 46529 (33.5) | 74.6 | 67.4 | 4.4 | 99.2 |
|  | >5 | 33822 (24.3) | 67.3 | 76.5 | 5.5 | 99.1 |
|  | >6 | 24126 (17.4) | 58.1 | 83.5 | 6.7 | 99.0 |
|  | >7 | 16867 (12.1) | 47.0 | 88.6 | 7.7 | 98.8 |
|  | >8 | 11669 (8.4) | 39.6 | 92.2 | 9.4 | 98.7 |
|  | >9 | 7939 (5.7) | 31.5 | 94.8 | 11.0 | 98.6 |
|  | >10 | 5123 (3.7) | 23.4 | 96.7 | 12.7 | 98.4 |
|  | >11 | 3103 (2.2) | 15.4 | 98.0 | 13.8 | 98.3 |
|  | >12 | 1884 (1.4) | 9.7 | 98.8 | 14.2 | 98.2 |
|  | >13 | 1249 (0.9) | 7.2 | 99.2 | 15.9 | 98.1 |
|  | >14 | 766 (0.6) | 4.7 | 99.5 | 16.8 | 98.1 |
|  | >15 | 428 (0.3) | 2.5 | 99.7 | 16.4 | 98.1 |
|  | >16 | 275 (0.2) | 1.4 | 99.8 | 14.2 | 98.0 |
|  | >17 | 133 (0.1) | 0.8 | 99.9 | 15.8 | 98.0 |
|  | >18 | 66 (0.0) | 0.4 | 100.0 | 16.7 | 98.0 |
|  | >19 | 21 (0.0) | 0.1 | 100.0 | 19.0 | 98.0 |
|  | >20 | 15 (0.0) | 0.1 | 100.0 | 26.7 | 98.0 |
|  | >21 | 2 (0.0) | 0.0 | 100.0 | 0.0 | 98.0 |
| WHO | >0 | 72599 (52.4) | 70.3 | 48.0 | 2.7 | 98.8 |
| TEWS | >0 | 124438 (90.9) | 96.5 | 9.3 | 2.1 | 99.2 |
|  | >1 | 63328 (46.2) | 77.6 | 54.4 | 3.3 | 99.2 |
|  | >2 | 39509 (28.8) | 63.9 | 71.9 | 4.4 | 99.0 |
|  | >3 | 18835 (13.8) | 44.9 | 86.9 | 6.4 | 98.7 |
|  | >4 | 8440 (6.2) | 27.9 | 94.3 | 8.9 | 98.5 |
|  | >5 | 3813 (2.8) | 18.2 | 97.5 | 12.9 | 98.3 |
|  | >6 | 1634 (1.2) | 10.2 | 99.0 | 16.8 | 98.2 |
|  | >7 | 708 (0.5) | 4.5 | 99.6 | 16.9 | 98.1 |
|  | >8 | 277 (0.2) | 2.2 | 99.8 | 20.9 | 98.1 |
|  | >9 | 84 (0.1) | 1.0 | 100.0 | 33.3 | 98.1 |
|  | >10 | 28 (0.0) | 0.2 | 100.0 | 21.4 | 98.0 |
|  | >11 | 8 (0.0) | 0.0 | 100.0 | 0.0 | 98.0 |
|  | >12 | 4 (0.0) | 0.0 | 100.0 | 0.0 | 98.0 |
| Quick COVID | >0 | 18914 (13.5) | 35.2 | 87.0 | 5.2 | 98.5 |
|  | >1 | 15026 (10.7) | 28.7 | 89.7 | 5.3 | 98.4 |
|  | >2 | 8973 (6.4) | 18.8 | 93.9 | 5.8 | 98.3 |
|  | >3 | 8210 (6.4) | 17.0 | 94.4 | 5.8 | 98.3 |
|  | >4 | 6301 (4.5) | 14.6 | 95.7 | 6.5 | 98.2 |
|  | >5 | 2560 (1.8) | 7.1 | 98.3 | 7.7 | 98.1 |
|  | >6 | 848 (0.6) | 2.9 | 99.4 | 9.6 | 98.1 |
|  | >7 | 305 (0.2) | 0.8 | 99.8 | 6.9 | 98.0 |
|  | >8 | 272 (0.2) | 0.7 | 99.8 | 7.0 | 98.0 |
|  | >9 | 85 (0.1) | 0.2 | 99.9 | 7.1 | 98.0 |
|  | >10 | 32 (0.0) | 0.2 | 100.0 | 18.8 | 98.0 |

**Supplementary Material 4: Performance of tools predicting composite secondary outcome death**
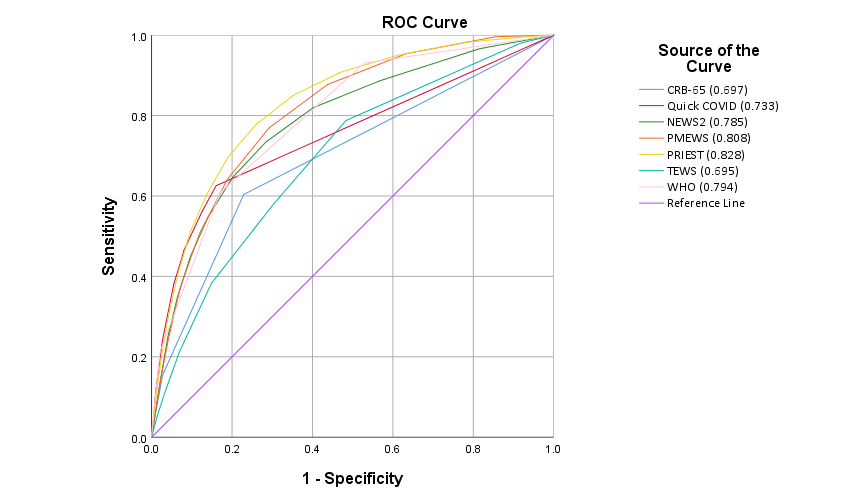

**Supplementary Material 5: Performance of tools predicting composite secondary outcome ICU admission**

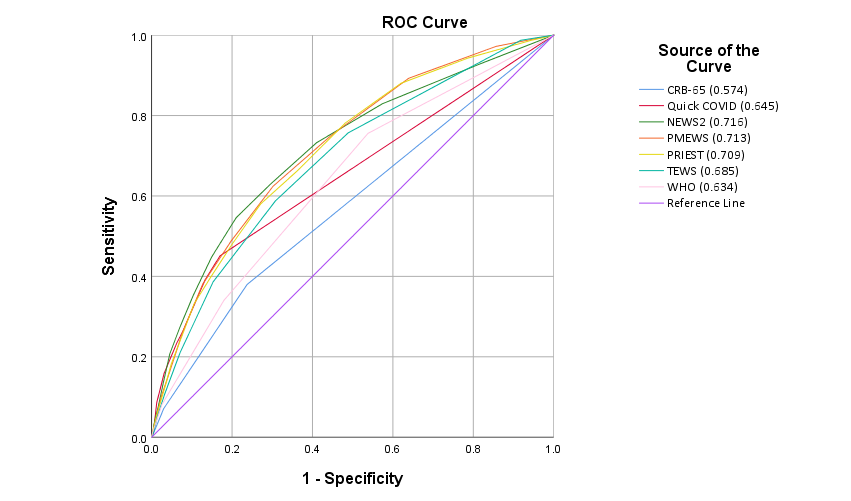

**Supplementary Material 6: Probability adverse outcome by score**

PRIEST

Probability adverse outcome (%)

| Score | Probability (%) |
| --- | --- |
| 0 | 0.24 |
| 1 | 0.37 |
| 2 | 0.7 |
| 3 | 1.17 |
| 4 | 1.85 |
| 5 | 2.49 |
| 6 | 3.94 |
| 7 | 6.1 |
| 8 | 7.47 |
| 9 | 10.53 |
| 10 | 13.02 |
| 11 | 15.95 |
| 12 | 18.26 |
| 13 | 20.04 |
| 14 | 23.39 |
| 15 | 25.65 |
| 16 | 27.35 |
| 17+ | 27.61 |

NEWS2

Probability adverse outcome (%)

| Score | Probability (%) |
| --- | --- |
| 0 | 0.66 |
| 1 | 0.92 |
| 2 | 1.43 |
| 3 | 2.36 |
| 4 | 3.14 |
| 5 | 5.37 |
| 6 | 8 |
| 7 | 9.69 |
| 8 | 12.76 |
| 9 | 16.35 |
| 10 | 18.28 |
| 11 | 19.89 |
| 12 | 20.99 |
| 13 | 24.46 |
| 14 | 22.9 |
| 15+ | 30.68 |

PMEWS

Proability adverse outcome (%)

| Score | Probability |
| --- | --- |
| 0 | 0.37 |
| 1 | 0.75 |
| 2 | 1.36 |
| 3 | 2.53 |
| 4 | 4.24 |
| 5 | 6.29 |
| 6 | 8.82 |
| 7 | 11.98 |
| 8 | 13.76 |
| 9 | 16.89 |
| 10 | 18.22 |
| 11 | 18.63 |
| 12 | 21.51 |
| 13 | 24.42 |
| 14 | 15.91 |
| 15+ | 29.01 |
